# Robust estimation of dynamic cerebrovascular reactivity using breath-holding fMRI: application in diabetes and hypertension

**DOI:** 10.1101/2023.05.20.23290209

**Authors:** Nuwan D. Nanayakkara, Liesel-Ann Meusel, Nicole D. Anderson, J. Jean Chen

## Abstract

Breath-holding (BH) tasks during functional magnetic resonance imaging (fMRI) acquisitions are gaining popularity for non-invasive mapping of carbon-dioxide (CO_2_) driven cerebrovascular reactivity (CVR), which is a valuable clinical marker of vascular function. However, compliance to BH tasks is often unclear, and the ability to record end-tidal CO_2_ often limited, rendering the optimal analysis of BH fMRI data a challenge. In this work, we demonstrate an adaptive data-driven approach for estimating CVR from BH fMRI data that minimizes errors due to subject non-compliance and regional CVR time delay variability. Building on previous work, we propose a frequency-domain-based approach for CVR estimation without the need for end-tidal CO_2_ (PETCO_2_) recordings. CVR amplitude is estimated in units of %ΔBOLD directly from the data-driven BH frequency. Serious deviations from the designed task paradigm were suppressed and thus did not bias the estimated CVR values. We demonstrate our method in detecting regional CVR amplitude and time-lag differences in a group of 56 individuals, consisting of healthy (CTL), hypertensive (HT) and diabetic-hypertensive (DM+HT) groups of similar ages and sex ratios. The CVR amplitude was lowest in HT+DM, and HT had a lower CVR amplitude than CTL regionally but the voxelwise comparison did not yield statistical significance. Notably, we demonstrate that the voxelwise CVR time delay estimated in Fourier domain is a more sensitive marker of vascular dysfunction than CVR amplitude. While HT+DM seems to confer longer CVR delays, HT seems to confer shorter delays than CTL. These are the first MRI-based observations of CVR time delay differences between diabetic-hypertensive patients and healthy controls. These results demonstrate the feasibility of extracting CVR amplitude and CVR time delay using BH challenges without PETCO_2_ recordings, and the unique clinical value of CVR time-delay information.

## 1. Introduction

Cerebrovascular reactivity (CVR) is a useful metric to assess how well the vasculature dilates or constricts in response to various stimuli. CVR is sensitive to various pathological conditions making it a potential biomarker [Giordani et al., 2014; Iranmahboob et al., 2016; Ivankovic et al., 2013; Liu et al., 2021; Petrica et al., 2007; Yazdani et al., 2020]. CVR also exhibits spatiotemporal variations even in the healthy brain due to regional differences in the cerebral vasculature [Bright et al., 2009; Lipp et al., 2015; Thomas et al., 2013]. Measurement of changes in CBF in response to a vasoactive stimulus is used to estimate CVR with the help of different neuroimaging methods such as transcranial Doppler ultrasound (TCD) [Kleiser and Widder, 1992], single-photon emission computed tomography (SPECT), and positron emission tomography (PET) [Ogasawara et al., 2003]. However, functional magnetic resonance imaging (fMRI) has been widely adopted for CVR measurements due to its ability to provide non-invasive whole-brain CVR estimation at an acceptable spatial and temporal resolution [Pinto et al., 2020; Urback et al., 2017]. The BOLD contrast is the most common for use in CVR mapping using MRI as it depends on the relative CBF changes in response to a vasoactive stimulus [Chen and Pike, 2010; Hauser et al., 2019; Mandell et al., 2008]. The advantages of direct measurement of CBF using non-invasive arterial spin labeling (ASL) perfusion imaging have also been demonstrated [Halani et al., 2015; Zhao et al., 2021], but the use of ASL is less frequent due to technical and availability constraints [Alsop et al., 2015; Heijtel et al., 2014].

Various vascular stimuli have been used to enable CVR mapping. The inhalation of air containing elevated CO_2_ content has been used frequently in CVR mapping [Liu et al., 2017b; Mark et al., 2010; Wise et al., 2007]. However, inhalation-based methods typically require complex experimental setups including breathing masks and are often not well tolerated by vulnerable groups such as children and the elderly [Spano et al., 2013]. Breath-hold (BH) tasks involve minimal setup and have been suggested as a robust alternative to gas challenges [Bright and Murphy, 2013; Lipp et al., 2015; Pinto et al., 2020; Urback et al., 2017]. BH tasks have been well tolerated by less cooperative subjects of vulnerable groups [Dlamini et al., 2018; Raut et al., 2016] and have been successfully applied in clinical studies [Haight et al., 2015; Iranmahboob et al., 2016; Raut et al., 2016; Tchistiakova et al., 2014]. A typical BH task follows a protocol consisting of a block design with alternating periods of normal breathing and breath holding for a simple reproducible voluntary breathing modulation task [Lipp et al., 2015; Peng et al., 2020; Pinto et al., 2016]. Nonetheless, well-known caveats of BH methods include the greater need for participant cooperation [Urback et al., 2017; Wu et al., 2015] and the unavailability of expired gas measurements during the BH periods, especially as a given BH task pattern across multiple subjects do not necessarily produce similar increases in CO_2_ partial pressure values in the blood [Urback et al., 2017] To help address this cause of uncertainty, some studies have incorporated measurement of end-tidal carbon dioxide partial pressure (PETCO_2_) using a mask or nasal cannula [Bright and Murphy, 2013; Pinto et al., 2016; Sousa et al., 2014]. A short BH period of 6 s can result in hypercapnia [Abbott et al., 2005] while the spatial extent of the significant BOLD response reaches a plateau at a BH length of around 20 s [Liu et al., 2002]. Many studies have reported a BH length of 15 s for successful estimation of CVR from BOLD data [Chang et al., 2008; Geranmayeh et al., 2015; Lipp et al., 2015; Tchistiakova et al., 2014; Urback et al., 2018]. With careful analysis, the performances of BH can be comparable to those involving gas manipulation techniques [Kannurpatti and Biswal, 2008; Liu et al., 2017a; Tancredi and Hoge, 2013; Wu et al., 2015]. However, at the CVR estimation step, uncertainties in subject compliance can present major data-analysis challenges, especially when PETCO_2_ recordings are unavailable.

Model-driven approaches such as the general linear model (GLM) have been employed to estimate CVR from BOLD responses to a BH task, whereby the BH paradigm is modelled as a boxcar [Biswal et al., 2007; Kastrup et al., 1999] or ramp function convolved with a hemodynamic response function (HRF) to approximate linear BOLD signal increases [Bright and Murphy, 2013]. Additional temporal derivatives of the HRF are required to account for the longer delays in respiratory responses to appear in the BOLD signal [Jahanian et al., 2017; Murphy et al., 2011]. The GLM model-driven approaches use a single fixed time lag in HRF to fully describe the BOLD signal across the whole brain. However, different brain regions exhibit different temporal CVR dynamics, both in healthy subjects and in patients [Chang et al., 2008; Geranmayeh et al., 2015; Magon et al., 2009; Pinto et al., 2016; Stringer et al., 2021]. Previous studies incorporating cross-correlations to estimate optimal time lags of the BOLD response have demonstrated substantial response-time variations across the brain [Chang et al., 2008; Geranmayeh et al., 2015]. The global BOLD time series has also been used in some studies as a reference to estimate the CVR time delay, ignoring the regional variations [Geranmayeh et al., 2015]. Iterative GLM fitting with shifted regressors [Geranmayeh et al., 2015; Moia et al., 2021; Murphy et al., 2011; Niftrik et al., 2016; Pinto et al., 2016] has also been successfully used in previous studies to estimate voxelwise CVR time delay. However, it is evident that GLM methods work best when PETCO_2_ recordings are available as input, allowing both inter-subject variations in BH compliance and in respiratory physiology to be accounted for [Birn et al., 2008; Bright and Murphy, 2013; Lipp et al., 2015; Murphy et al., 2011]. Moreover, the canonical hemodynamic response function is derived using the BOLD response to neuronal activity and not CO_2_.

We propose a novel approach for estimating the amplitude and timing of the BH BOLD response without modelling or correlation with other signals (e.g. PETCO2). Our approach makes use of the Fourier representation of the spectrum of BOLD data, as it has been found that the BH CVR response can be successfully approximated as a sinusoidal signal by assuming the BH task is approximately symmetrical (equivalent BH and baseline periods), even if it is not [Lipp et al., 2015; Murphy et al., 2011]. This sinusoidal approach was shown to outperform the use of an HRF-based regressor (the PETCO_2_ trace convolved with the HRF [Niftrik et al., 2016]), prompting us to estimate CVR in the Fourier domain. Indeed, in the Fourier-series-based regression approach by Pinto et al., BH designs that deviate further from symmetry can be addressed by adding higher harmonics to the Fourier series [Pinto et al., 2016]. On the other hand, our data-driven Fourier-based approach does not require regressors but instead detects and accounts for deviations from task designs directly from the BOLD fMRI data. In this study, we used a typical BH task design similar to the studies mentioned earlier. We demonstrate our method by assessing differences in CVR amplitude and time delay among patients with hypertension (HT), hypertension-plus-type-2 diabetes (HT+DM), and age-matched controls (CTL).

## 2. Methods

### 2.1. Study participants

Older adults of ages 65-85 were recruited and placed in control (CTL), hypertension (HT), or hypertension-plus-type-2 diabetes (HT+DM) groups on the basis of screening measures. Participants were excluded from the study if they met any of the following criteria: (1) a score ≤ 31 on the Telephone Interview for Cognitive Status – modified version [Welsh et al., 1993] in order to exclude participants with possible dementia; (2) the use of insulin to treat DM; (3) the presence of DM complications, based on self-report, including clinically significant gastroparesis, retinopathy, nephropathy, neuropathy, hepatic disease, or a recent coronary heart disease event as determined by a physician; (4) other significant medical or psychiatric disorders affecting cognitive function, such as stroke (self-report or evidence from structural scans) and major depressive disorder; (5) current or recent use of central nervous system-active medications, including those for the treatment of depression, sleep disorders, and migraine headaches; (6) major inflammatory disorders, heart failure, and chronic lung disease; or (7) hormone replacement therapy in female participants. The included participants were screened to ensure group status as listed below:

- CTL: Participants had a mean systolic BP ≤ 140 mmHg, a mean diastolic BP ≤ 90 mmHg, no history of antihypertensive medication use, and a fasting glucose level (FGL) ≤ 6.1 mmol/L.
- HT: Participants were using antihypertensive medication under physician prescription for a minimum of two years, with current blood pressure within a normal or HT range and limited to those who were using long-acting antihypertensive medications (e.g., ACEIs, angiotensin II receptor blockers, diuretics) in order to capture the most commonly prescribed medications.
- HT+DM: Participants had a physician diagnosis of type 2 DM for a duration of at least two years, were controlling their DM through diet or hypoglycemic medication alone, and were free of major DM complications as defined in the exclusion criteria, in addition to the criteria for the HT.

All selected participants completed a brief medical questionnaire, which included questions about the use and duration of all medications and the duration of HT and DM. Participants provided a fasting blood sample for measurement of hematocrit, lipid profile (triacylglycerides [TG], total cholesterol [TC], low-density lipoprotein [LDL], and high-density lipoprotein [HDL]), CRP, glucose, insulin, and HbA1c. Blood pressure, weight, height, and waist circumference were also measured. These measurements were followed by a practice session of the breath hold task in an MRI simulator to ensure that the participant was comfortable with the fMRI scanning protocol. Participants were asked to continue their usual diet, medications, and activity level for the duration of their involvement in the study. Available data from 56 participants (CTL:21, HT: 23, and HT+DM: 12) were preprocessed as described in Section 2.3. Data from seven participants (CTL:3, HT:3, and HT+DM:1) were removed from analysis due to excessive head-motion artifacts larger than 1⁰ rotation and 1 mm translation as detected by FSL motion correction. This study consisted of remaining data from 49 participants (CTL: 18, HT: 20, and HT+DM: 11) with mean ages of 70.2 ± 3.3, 71.9 ± 4.7, and 71.7 ± 3.6 years, and male/female ratios of 1.25, 0.43 and 0.57, respectively.

### 2.2. Data acquisition

Each participant followed a set of 6 repetitions of a 30 s resting and 2 s exhale followed by a 15 s BH guided by visual clues (total duration, *T* = 47 s) during a dual-echo pCASL fMRI image acquisition session on a Siemens Trio 3T system (T2*-weighted echo-planar imaging, FOV = 220 mm, acquisition matrix = 220 x 220, voxel size = 3.4 x 3.4 x 6.0 mm, bandwidth = 2790 Hz/Pixel, TE1/TE2/TR = 9.1/25/4000 ms, flip angle = 90 degrees, slices = 16, averages = 1, concatenations = 1, scan duration = 5:24). The data associated with the second TE were used to compute the BOLD time series. Respiratory bellows were recorded using the scanner’s built-in belt. A T1 anatomical scan (FOV = 256 mm, acquisition matrix = 192 x 256, voxel size = 1.0 mm^3^, bandwidth = 200 Hz/Pixel, TI/TE/TR = 1100/2.63/2000 ms, flip angle = 9 degrees, slices = 160, averages = 1, concatenations = 1, scan duration = 6:26) was acquired for anatomical reference and tissue segmentation.

### 2.3. Preprocessing

The dual-echo pCASL time series data were preprocessed using FSL [Jenkinson et al., 2012; Smith et al., 2004] and AFNI [Cox, 1996; Cox and Hyde, 1997] tools. Slice timing correction using *slicetimer* in FSL was applied to correct for sampling offsets inherent in slice-wise EPI acquisition sequences. The BOLD data in the dual-echo pCASL time series were isolated from the full pCASL time series and separated into odd and even time frames. Both of these time series were separately corrected for subject motion using the *mcflirt* and registered into MNI space using *flirt*. AFNI’s *3dretroicor* was used to generate BOLD data that were corrected for noise associated with physiological motion (i.e., heartbeat, respiration). Neighbouring control and tag frames were averaged in a sliding-window manner to produce 78 frames of BOLD data, which were then high-pass filtered to remove low-frequency noise from the data with a 0.01 Hz cut-off frequency, resulting in the “preprocessed BOLD signal”.

### 2.4. Data-driven CVR estimation

CVR estimation was performed on the preprocessed BOLD data using our robust data-driven pipeline shown in **Figure 1**. The BOLD data was first normalized by the mean of the first 8 frames of the series corresponding to the baseline before the first BH period. The time series data was then converted to %ΔBOLD by voxelwise demeaning. As described earlier, the identification of the BH frequency (BHF) can be hampered by variations in task compliance as well as in participant physiology. Thus, the identification of the BHF in our pipeline undergoes a two-step process to ensure flexibility and robustness. First, we identified the BH task paradigm used in the study that can be considered as a repetitive signal of period *T* = 47 s, corresponding to a fundamental BHF of 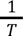 (0.0213 Hz). The voxelwise BOLD signal was passed through a bandpass filter of cut-off frequencies 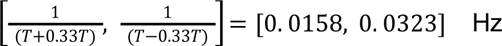, corresponding to *T*±*T/3*. The pipeline finds the BHF from the BOLD data spectrum within this frequency range, such that the BHF can deviate slightly from the nominal frequency due to variations in subject compliance with the task design, but is not allowed to deviate too far away from the expected task frequency to ensure robustness against noise. Second, the pipeline finds the BHF corresponding to the maximum amplitude of the Fourier spectrum of the BOLD signal at each cortical voxel. Lastly, for the grey matter (GM) of each data set, a histogram is constructed using the voxel-wise BHF, and the BHF found at the peak occurrence is chosen as the dominant BHF for that data set. This second step further constrains the BHF to maximize inter-regional comparability, assuming that variations in respiratory physiology contribute to inter-subject but not within-subject inter-regional CVR differences. That is, the dominant BHF can vary between participants but not between brain regions.

**Figure 1.**
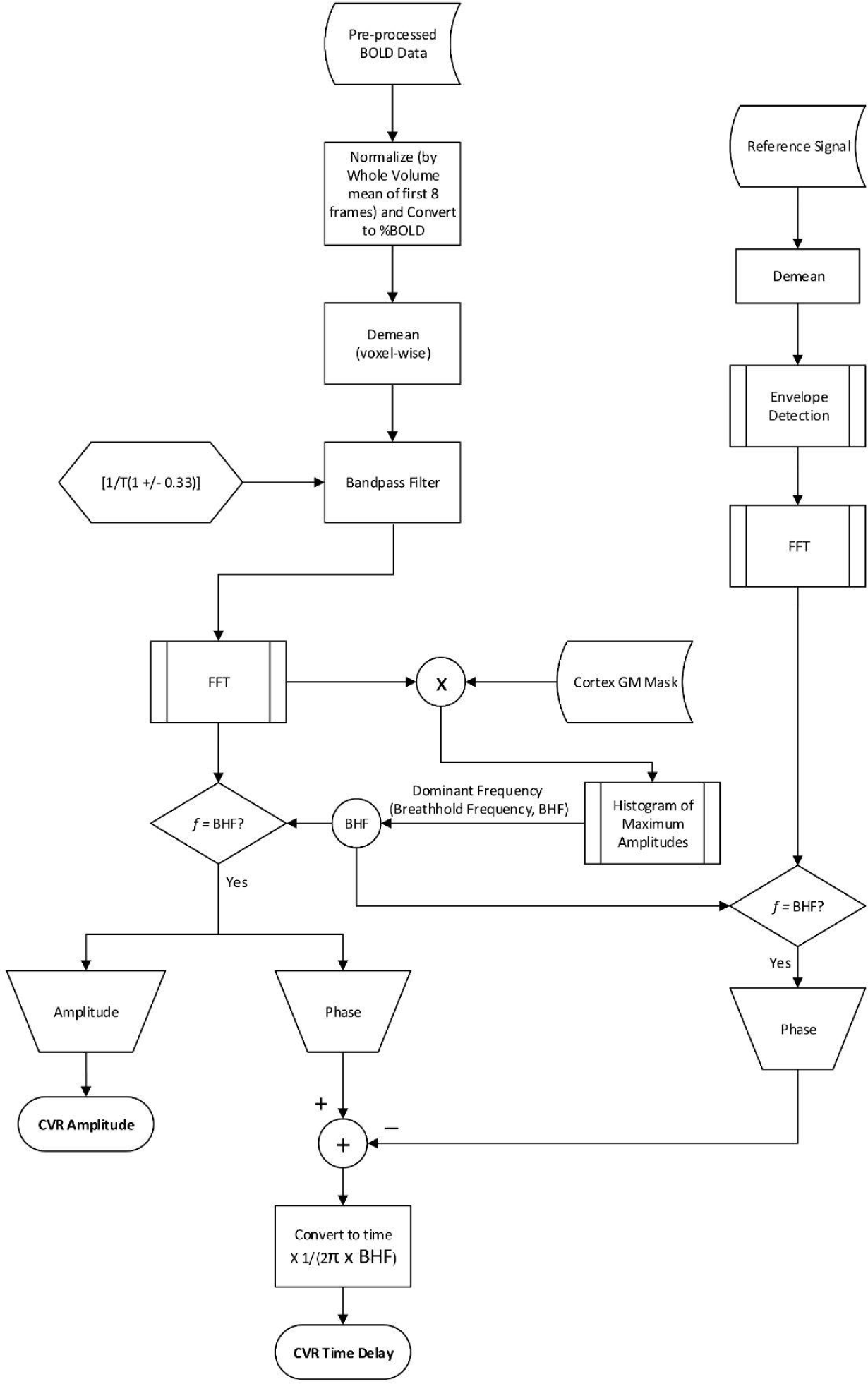
Signal processing pipeline. The normalized and demeaned %ΔBOLD data were bandpass filtered to retain signals around the targeted BH frequency (BHF). The dominant frequency of the filtered signal in the cortex was selected as the BHF. The amplitude of the spectrum of each voxel the BHF was selected as the CVR amplitude. The CVR response time of each voxel was calculated relative to the BH task extracted at the BHF using the envelope of the respiratory belt signal.

When a reference signal is available, the voxelwise phase (ϕ_*s*_) of the CVR at BHF relative to a reference signal at the same frequency can be used to compute a response lag for the CVR response. One such reference is the respiratory belt recording, which was used in this study. The envelope of the respiratory signal was extracted from the recording, which also allowed us to verify each subject’s compliance with the BH task. The phase (ϕ_*r*_) of the de-meaned respiratory belt signal envelope Fourier spectrum at the BHF was selected as the reference for CVR time delay, calculations as

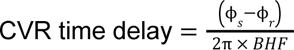

### 2.5. Statistical analysis

We also computed the Framingham Risk Score (FRS) [Wilson et al., 1998] and Diabetes Epidemiology: Collaborative Analysis of Diagnostic Criteria in Europe (DECODE) scores [Balkau et al., 2004]. Differences in demographics and physiological measurements across groups, including the FRS and DECODE metrics, were detected by ordinary one-way ANOVA corrected for multiple comparisons by controlling the false discovery rate using the Benjamini-Hochberg procedure.

The CVR amplitude and time delay were compared voxelwise and region-wise across the three groups (namely CTL, HT, and HT+DM). The GLM approach [Winkler et al., 2014] was used for the voxelwise comparison followed by multiple comparisons correction via threshold-free cluster enhancement [Smith and Nichols, 2009]. Brain parcellations generated by nonlinear spatial registration of T1-anatomical scan to the default GCA atlas in FreeSurfer (Version 6.0.1, available at surfer.nmr.mgh.harvard.edu) were used to calculate mean CVR amplitudes and mean CVR time delays in cortical regions in each group. The mean regional CVR amplitudes were normally distributed in all three groups while the distributions of time delays were not normal when tested using the D’Agostino-Pearson test [D’Agostino and Stephens, 1986] for normality (see supplementary table S1). Two-way ANOVA with multiple comparisons was used to compare the overall means of region-wise (ROI) CVR amplitude, corrected for multiple comparisons by controlling the false discovery rate (Benjamini-Hochberg procedure) between groups. The statistically significant differences detected by Friedman’s test were corrected for multiple comparisons by controlling the false discovery rate using the Benjamini-Hochberg procedure for whole group-wise comparison of overall means of CVR time delay. Regional CVR amplitude and time delay were compared seperately in each cortical region of interest (ROI) using the Kruskal-Wallis test corrected for multiple comparisons by controlling the false discovery rate.

## 3. Results

Figure 2 summarizes the demographics and physiological measurements for participants in the three groups. Participant age was not significantly different across the groups (**Figure 2a**). HT and HT+DM groups did not exhibit significant differences in systolic BP (SBP) compared to CTL (**Figure 2b**). The HT+DM group showed significantly higher HbA1C and lower LDL cholesterol values compared to the HT and CTL groups (**Figure 2c, d)**. The FRS and DECODE scores were both significantly lower in CTL compared to HT and HT+DM, but only the DECODE score showed a significantly lower score for HT compared to HT+DM (**Figure 2e, f**).

**Figure 2.**
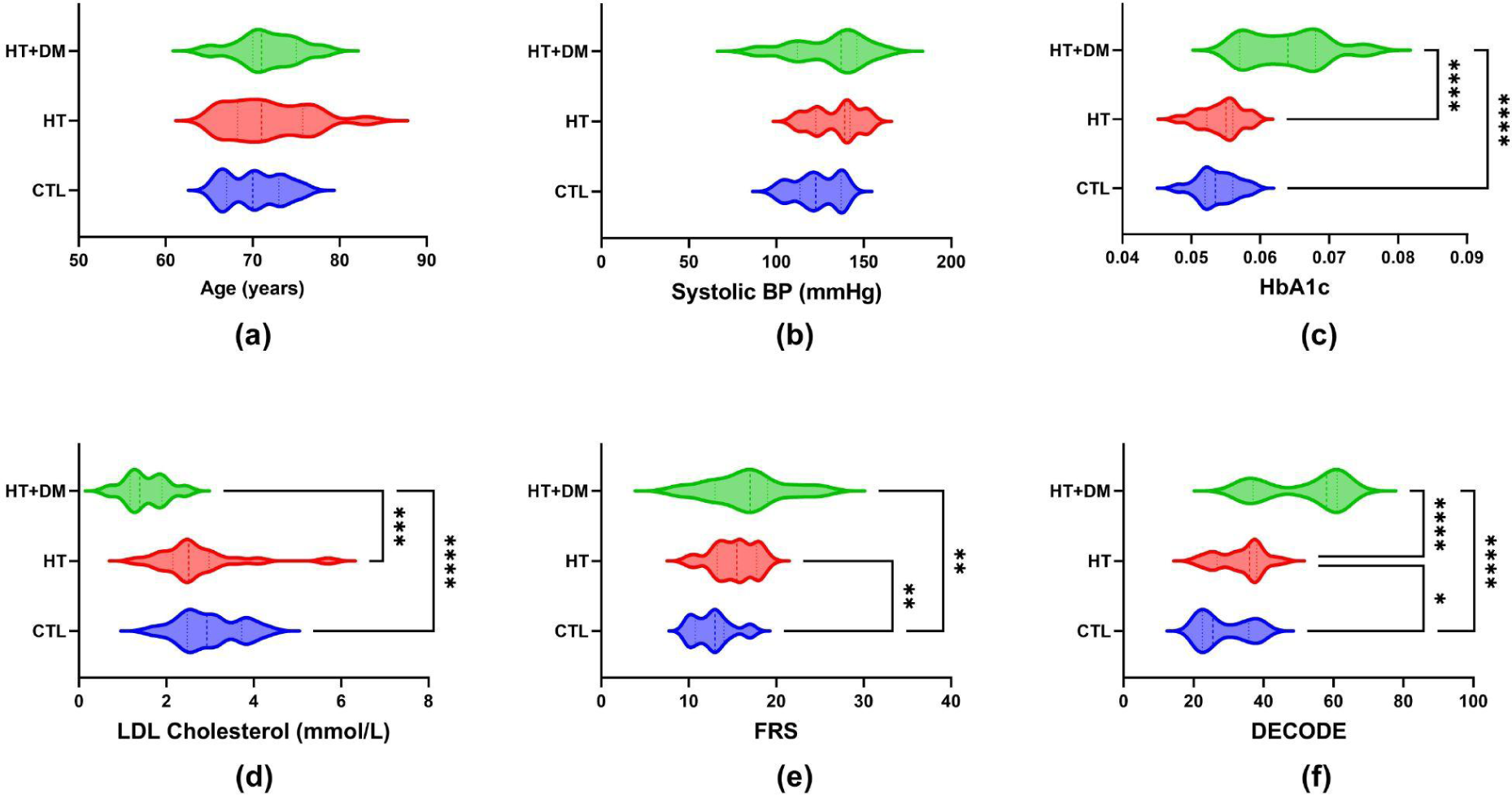
Subject demographics. The mean (a) age, (b) systolic blood pressure, (c) HbA1c values, (d) LDL cholesterol, (e) Framingham Risk Score (FRS), and (f) Diabetes Epidemiology: Collaborative Analysis of Diagnostic Criteria in Europe (DECODE) score for each subject group. The statistically significant differences were detected by ordinary one-way ANOVA corrected for multiple comparisons by controlling the false discovery rate using the Benjamini-Hochberg procedure indicated by * (q < 0.05), ** (q < 0.01), *** (q < 0.001) and **** (q < 0.0001).

Shown in **Figure 3** are sample signals from the intermediate steps of the signal processing pipeline for a participant compliant (3a) and a participant non-compliant (3b) to the BH task design. Bandpass filtering suppressed spurious signals outside the targeted BHF range, as in Figure 3a(ii) and 3b(ii). The histograms of maximum amplitudes of the bandpass-filtered BOLD spectra in all cortical voxels are shown in **Figure 3a(i) and 3b(i)** for sample subjects. The peaks of the histograms correspond to the dominant CVR frequencies. In addition, the BOLD signal at the dominant frequency is well-modelled by the sinusoidal approximation and closely follows the respiratory belt signal as shown in **Figure 3a(v) and 3b(v)**, even when the subject performance on the BH task deviated from the task design.

**Figure 3.**
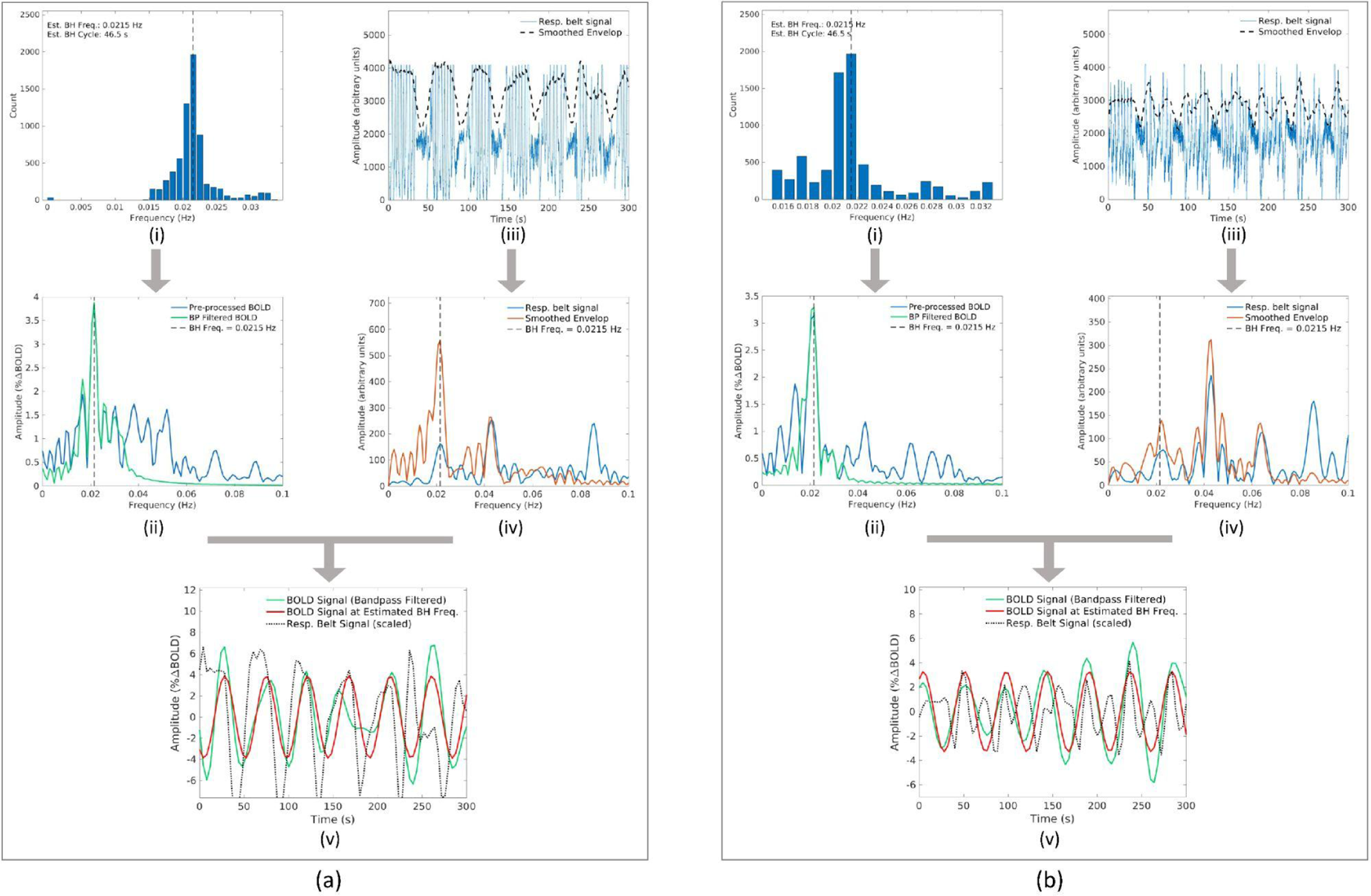
Samples outputs from intermediate signal-processing steps. Samples from the signal processing pipeline for a subject (a) compliant and (b) non-compliant to BH task design. Each includes (i) the histogram of maximum amplitudes at all cortical voxels with corresponding frequencies, (ii) the frequency spectrum of the pre-processed and band-pass filtered BOLD at a sample voxel in the cortex, (iii) the respiratory belt signal with the smoothened envelope, (iv) respective frequency spectrum of the belt signal, and (v) bandpass filtered the BOLD signal, and the estimated CVR response at the BHF at the sample voxel in the cortex.

**Figure 3** shows the recorded full respiratory-belt signal and the extracted BH pattern from the smoothed envelope details with corresponding frequency spectrums for a participant who followed the BH task paradigm (**figures 3a(iii)** and **3a(iv)** respectively) and for a participant who was not compliant (**figures 3b(iii)** and **3b(iv)** respectively). As indicated by dashed lines on respective frequency spectrums, the proposed data-driven approach was able to correctly estimate the BHF in both cases avoiding peaks from unexpected frequencies.

**Figure 4** shows the mean CVR amplitude maps in each group. GM regions in the cortex generally have a higher CVR amplitude compared to other areas of the brain. The mean CVR amplitude in CTL was significantly higher than both HT and HT+DM across cortical GM and higher in HT compared to HT+DM (see supplementary figure S1a).

**Figure 4.**
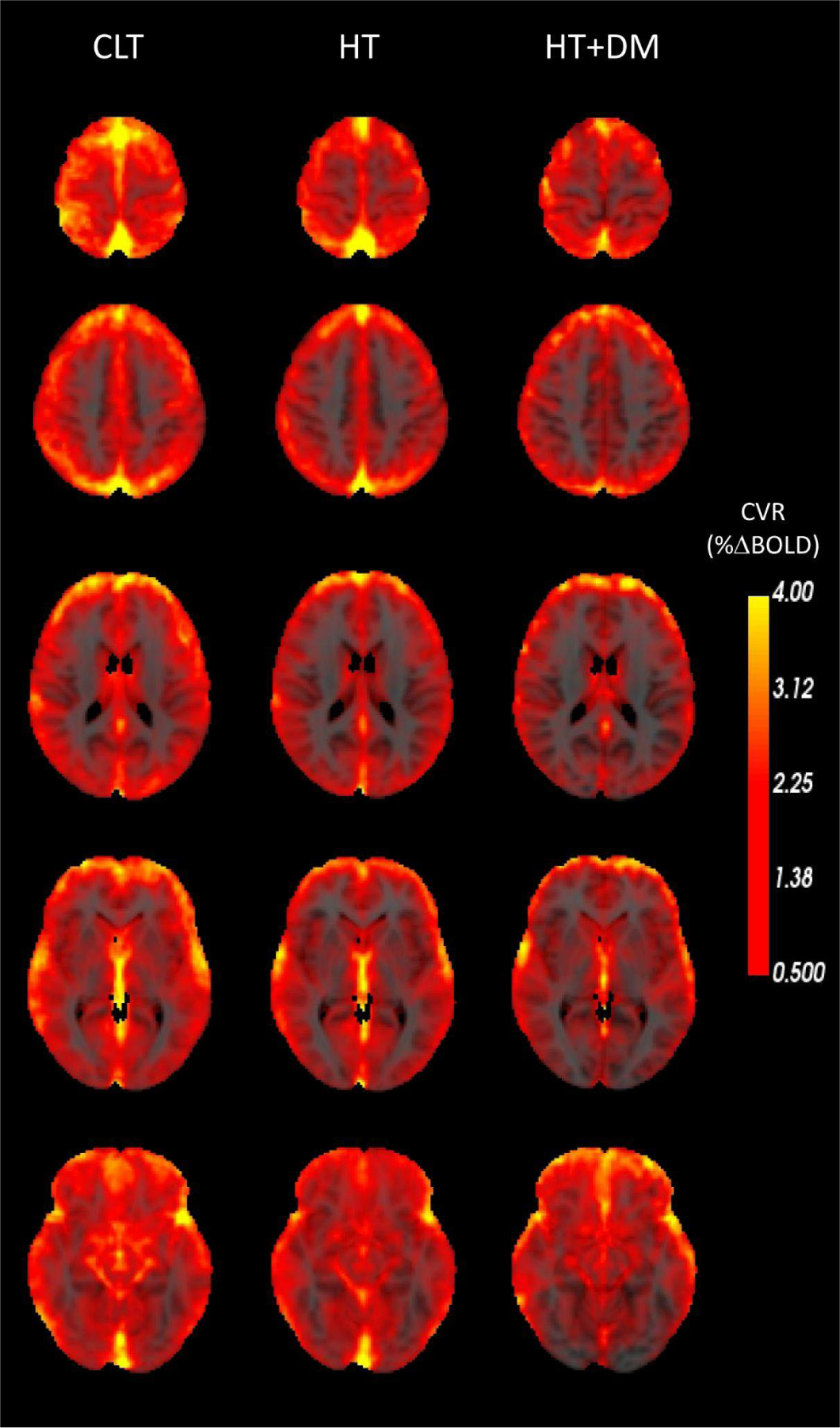
Mean CVR amplitude (%ΔBOLD) for all subjects in each group. The grey matter (GM) regions in the cortex have a higher CVR amplitude than the white-matter (WM) and other areas of the brain. The HT+DM participants showed the lowest CVR amplitudes consistently throughout all brain regions.

**Figure 5** shows the mean CVR time delay maps in each group. GM regions in the cortex generally have a shorter delay than in other areas of the brain. The mean CVR time delay in HT+DM was significantly longer than both CTL and HT across cortical GM and longer in CTL compared to HT (see supplementary **figure S1b**).

**Figure 5.**
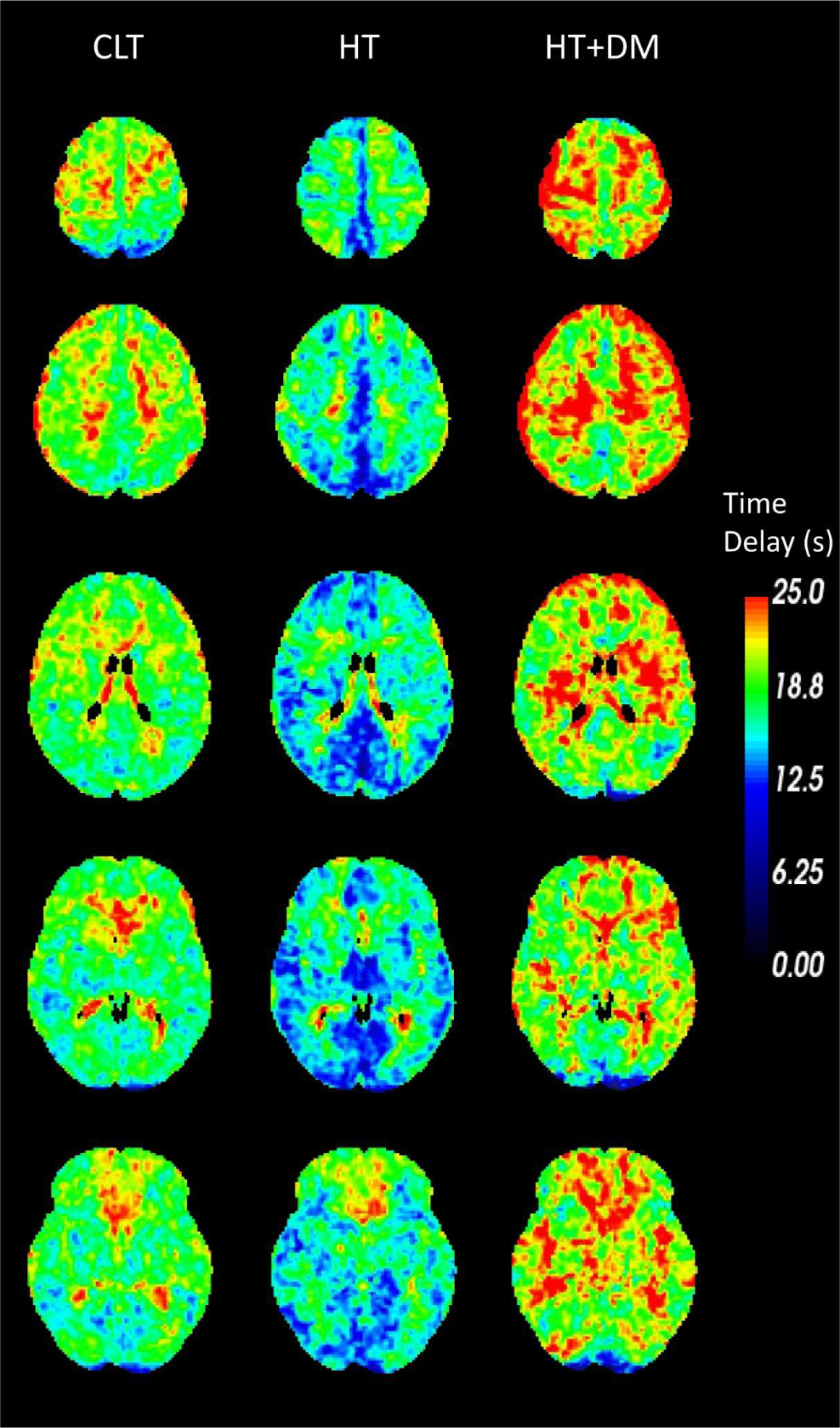
Mean CVR time delay (s) relative to the respiratory belt signal in each group. The GM regions in the cortex have a shorter CVR time delay than the WM and other areas of the brain. The HT group showed the lowest CVR time delay while HT+DM group showed the longest delay.

**Figure 6** shows voxels that demonstrate statistically significant differences (p < 0.05) where CVR amplitudes in CTL > HT+DM. Most CVR amplitude differences were reduced when controlling for sex, the duration of previously detected hypercholesterolemia, or systolic blood pressure. No statistical significance voxels were detected when controlling for LDL or HbA1c. No significant voxelwise differences in CVR amplitudes were detected between CTL and HT or between HT and HT+DM.

**Figure 6.**
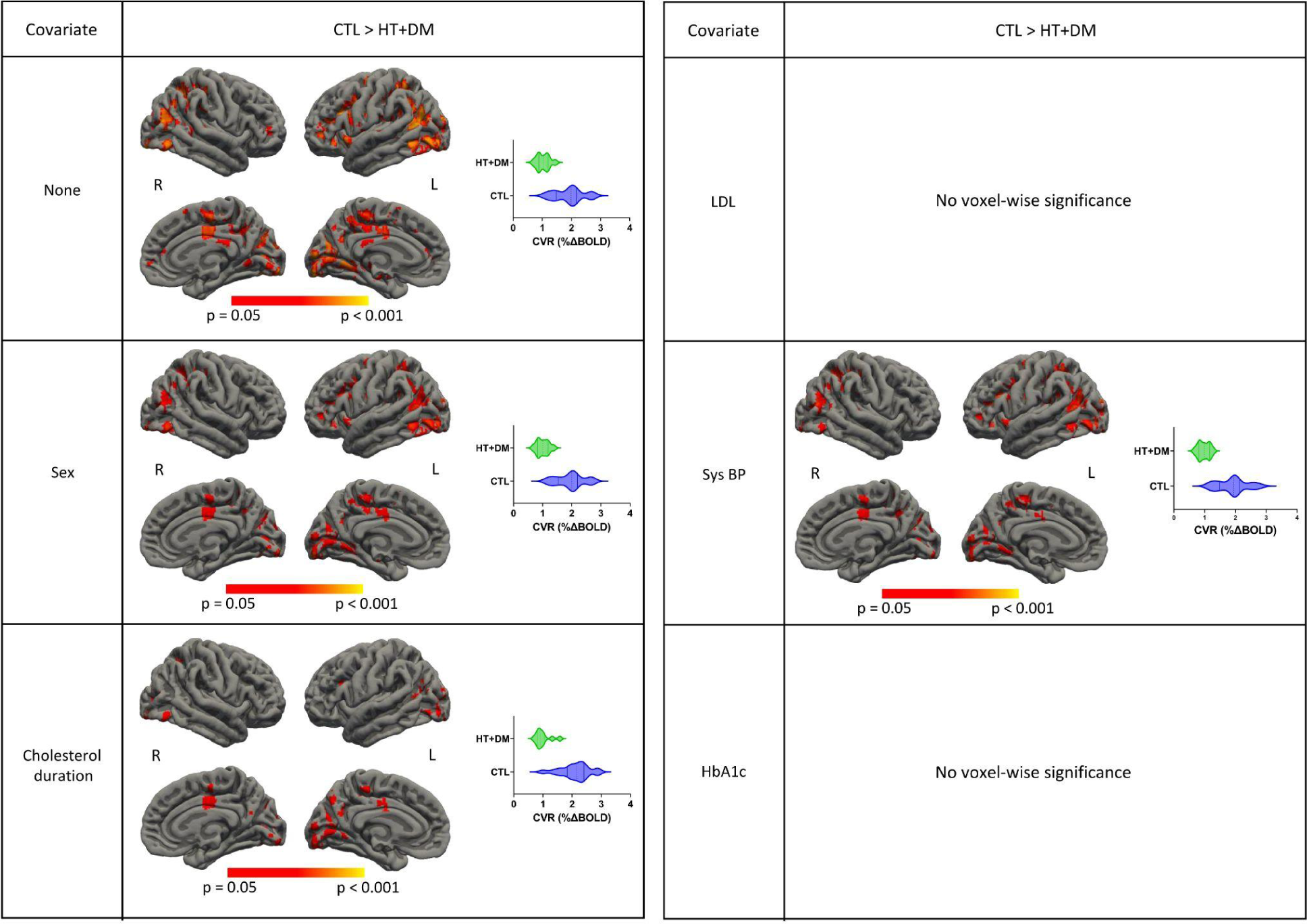
Voxelwise comparisons of CVR amplitudes. The statistical testing on CVR amplitudes of CTL > HT+DM showed significant voxels (p < 0.05) for permutation inference for the general linear model after threshold-free cluster enhancement. The significance of differences in most voxels was reduced when controlled for sex, the duration of previously detected hypercholesterolemia, or systolic blood pressure. No statistical significance voxels were detected when controlling for LDL or HbA1c. Significant differences in CVR amplitude were not detected between CTL and HT or between HT and HT+DM.

**Figure 7** shows voxels that demonstrate statistically significant differences (p < 0.05) where CVR time delays in CTL > HT and HT < HT+DM. More significant differences were detected for HT < HT+DM than CTL > HT and those differences were reduced when controlling for the duration of previously detected hypercholesterolemia and disappeared when controlling for LDL and HbA1c. Significant differences in CVR time delay were not detected between CTL and HT+DM.

**Figure 7.**
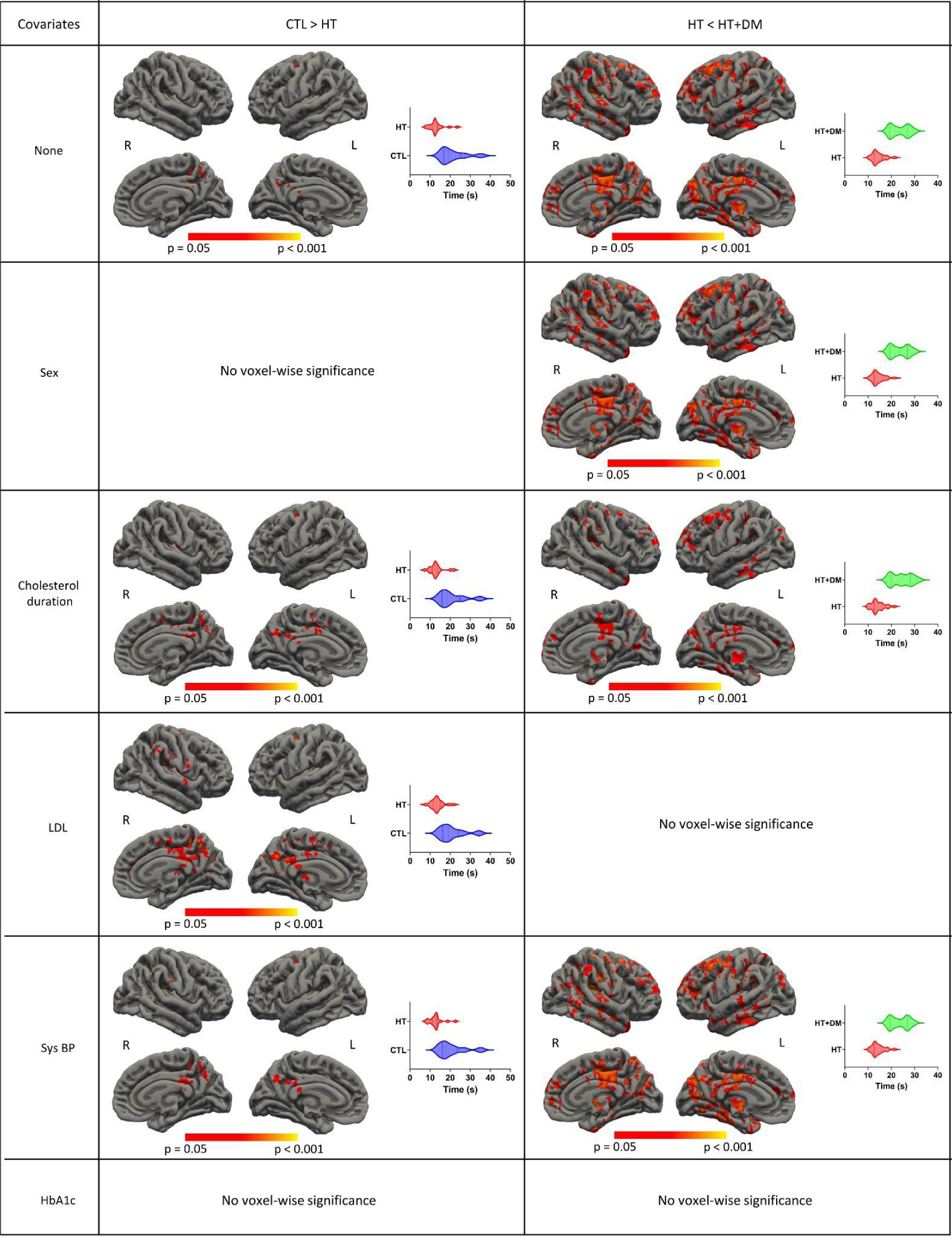
Voxelwise comparisons of CVR time delay. The statistical testing on the CVR time delay of CTL > HT and HT < HT+DM showed significant voxels (p < 0.05) for permutation inference for the general linear model after threshold-free cluster enhancement. More significant differences were detected for HT < HT+DM than CTL > HT; those were reduced when controlling for the duration of previously detected hypercholesterolemia and disappeared when controlling for LDL and HbA1c. Significant differences in CVR time delay were not detected between CTL and HT+DM.

**Figure 8** shows the regional means of CVR amplitude for each cortical ROI. CVR amplitude in HT+DM is significantly lower than both CTL and HT in the cuneus, the lingual, the precuneus, the isthmus cingulate, the lateral occipital, the pericalcarine, and significantly lower than CTL in the posterior cingulate.

**Figure 8.**
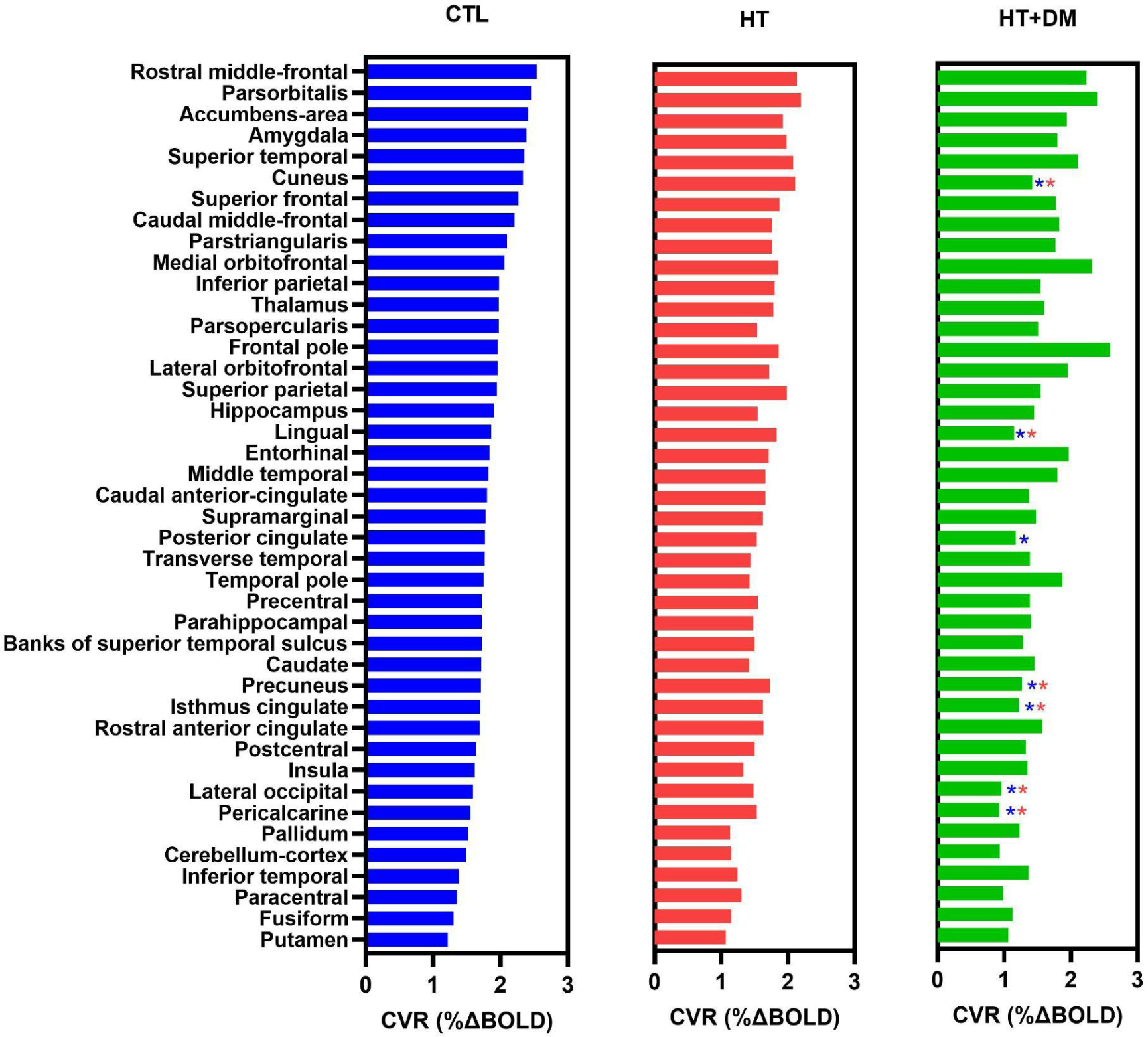
Mean regional CVR amplitude arranged in the descending order of CTL in subregions of the cortex. CTL showed the highest amplitude and HT+DM showed the lowest with a few exceptions. HT+DM showed statistically significant lower CVR amplitudes (p < 0.05) in multiple cortical regions from both CTL and HT with the Kruskal-Wallis test corrected for multiple comparisons by controlling the false discovery rate as marked by blue (*) and red (*) asterisks, respectively.

**Figure 9** shows the regional means of CVR time delay for each cortical ROI. HT exhibited significantly shorter delays than CTL in the cuneus, the lingual, the isthmus cingulate, the superior frontal, the posterior cingulate, the thalamus, the pallidum, the rostral anterior cingulate, and the frontal pole. The CVR time delay was significantly longer in HT+DM than in both CTL and HT in the entorhinal, the superior parietal, the inferior temporal, the fusiform, the postcentral, the banks of the superior temporal sulcus, the middle temporal, the superior temporal, the lateral orbitofrontal, and the posterior cingulate. HT+DM also showed a significantly longer CVR time delay in many regions compared to HT.

**Figure 9.**
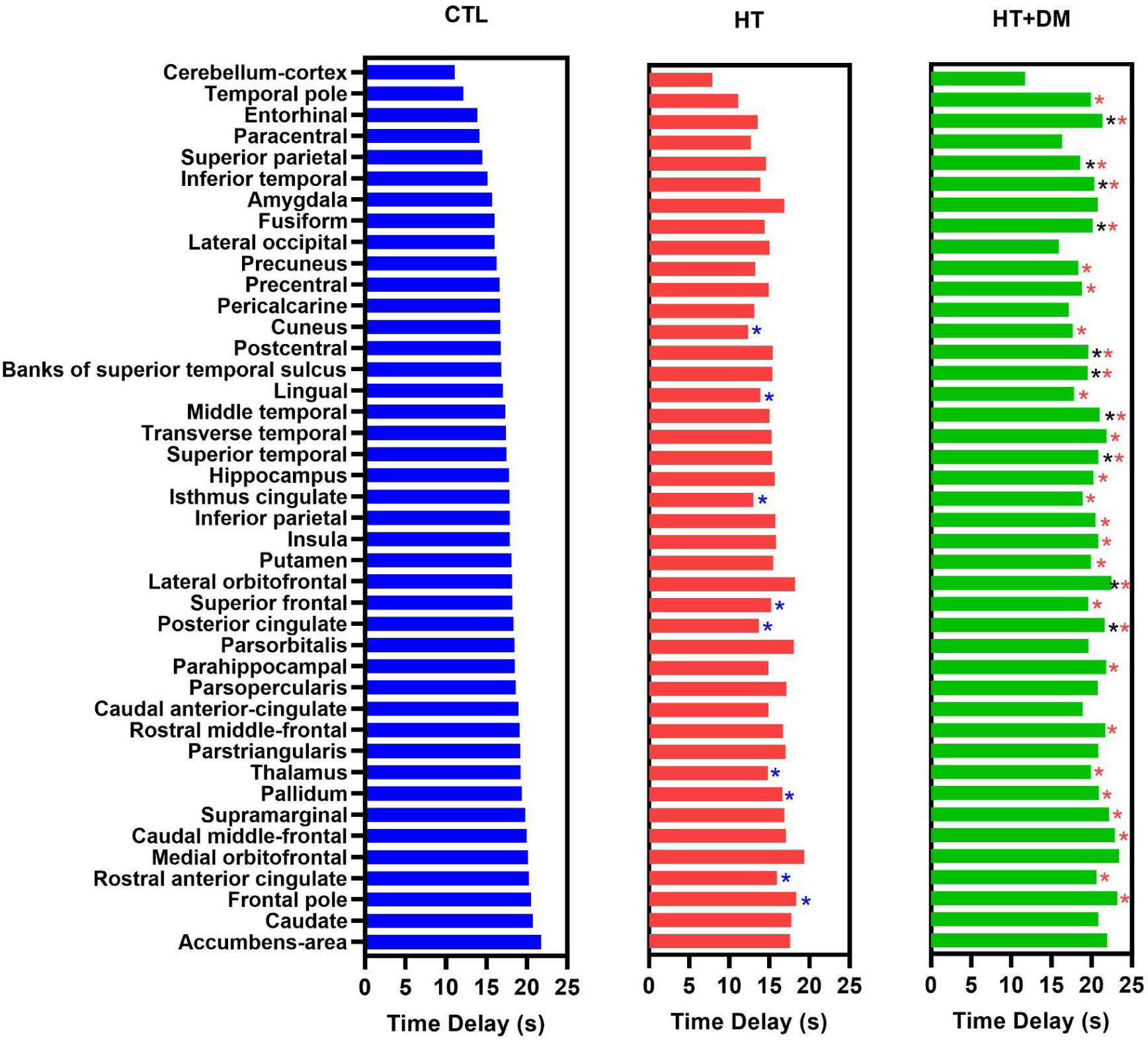
Mean regional CVR time delay arranged in the ascending order of CTL in subregions of the cortex. HT group showed the fastest CVR response and HT+DM showed the slowest. The CVR time delay showed statistically significant differences in more regions than those for amplitude with the Kruskal-Wallis test corrected for multiple comparisons by controlling the false discovery rate. Statistically significant shorter CVR time delay compared to CTL is marked with blue (*) and longer CVR time delays compared to CTL and HT are marked by black (*) and red (*) asterisks, respectively.

## 4. Discussion

In this work, we present a simple, frequency-domain-based, data-driven approach for estimating CVR from BH fMRI data. Our adaptive, data-driven approach for CVR estimation helps to prevent errors due to subject non-compliance and regional CVR time delay variability. It thus can have a wide range of applications in studying patient populations. Moreover, our Fourier-spectrum approach provides an elegant means to estimate voxelwise CVR time delay estimations using phase differences relative to any non-neural biological signal that depends on the BH task [Blockley et al., 2011; Pinto et al., 2020]. We demonstrate our method in the study of diabetes and hypertension.

### 4.1. Estimation and interpretation of CVR amplitude using BH

A BH task is a simple method to induce cerebrovascular response due to an increase of arterial CO_2_ levels by ceasing ventilation. It provides a reproducible technique estimation of CVR even in less cooperative populations [Bright et al., 2009; Lipp et al., 2015; Peng et al., 2020]. The BH-induced CVR has been shown to be comparable to CVR estimated with inhaled CO_2_ challenges [Chan et al., 2020; Kastrup et al., 2001; Prakash et al., 2014; Raut et al., 2016]. Model-driven approaches, mainly the general linear model (GLM), are being used for estimating CVR from BOLD data acquired during BH tasks. A function describing the block design of the breathing paradigms convolved with the hemodynamic response function is typically used to build these models of the BH CVR response [Biswal et al., 2007; Kastrup et al., 1999]. The GLM models perform well when PETCO_2_ recordings are available for modelling BOLD response to accommodate for inter- and intra-subject variations [Bright and Murphy, 2013; Pinto et al., 2016]. However, the performance of GLM methods on data from non-compliant subjects is questionable [Jahanian et al., 2017]. The BOLD fMRI signal changes due to head motion, confounding physiological fluctuations, and other sources of noise can affect the CVR estimation from BH BOLD data [Chen et al., 2021; Jorge et al., 2013; Moia et al., 2021]. Head motion can be particularly problematic in BH CVR estimation [Johnstone et al., 2006; Moia et al., 2021]. The data affected by extensive head motion were excluded from this study instead of using complex motion correction algorithms while using the standard motion correction algorithms for correcting smaller motions. Respiration can also disturb the B0 field due to the change of air in the lungs [Raj et al., 2001] and introduce aliasing artefacts or pseudo-movement effects in the signal [Pais-Roldán et al., 2018; Power et al., 2019]. Such effects can easily result in vulnerability of the BH data analysis approach.

Alternatively, Fourier basis modelling has been used effectively, assuming sinusoidal signal variations at the paradigm design frequency [Bright and Murphy, 2013; Murphy et al., 2011; Pinto et al., 2016]. Fourier-based approaches have been reported as being robust and versatile, but are seemingly more ideal for BH designs that are symmetrical. In practice, the BOLD response to BH tasks can easily deviate from the sinusoidal frequency, and it is unclear what the implications are for CVR estimation.

In our data-driven approach, we addressed this issue by estimating fundamental frequency from the BOLD signal spectrum for accurate sinusoidal approximation of CVR. GM regions in the cortex generally have a higher CVR amplitude (**Fig. 4**) than the WM and other areas of the brain. Regional variations within the GM are also visible. Frontal regions generally have a higher CVR amplitude compared to temporal regions, the cerebellum cortex, the fusiform, and the putamen. These overall and regional differences are consistent with previous similar studies [DuBose et al., 2022; Moia et al., 2021; Thrippleton et al., 2018], and attest to the robustness of our simple approach.

### 4.2. CVR amplitude in diabetes and hypertension

Cerebrovascular reactivity (CVR) changes have been reported in various chronic conditions [Atwi et al., 2019; Geranmayeh et al., 2015; Iranmahboob et al., 2016; Ivankovic et al., 2013; Li et al., 2021; Yazdani et al., 2020]. Chronic hypertension [Harvey et al., 2015] has been associated with CVR impairments [Ivankovic et al., 2013; Kadoi et al., 2003; Li et al., 2021; Petrica et al., 2007; Yazdani et al., 2020]. Specifically, in HT, hypercapnia-based BOLD MRI uncovered extensive reductions in blood flow and CVR in hypertensive rats compared to controls [Li et al., 2021]. In older adults, HT is well known to be associated with arterial stiffness and blood-flow reduction [Tomoto et al., 2023] in addition to reductions in whole-brain CVR [Jefferson et al., 2018]. This CVR reduction is consistent with reduced resting-state fMRI signal fluctuations in the presence of arterial stiffness, predominantly in the precuneus, anterior and posterior cingulate gyrus, paracingulate gyrus, and frontal-pole regions [Hussein et al., 2020]. The reduced CVR is in turn associated with impaired executive function [Hajjar et al., 2014], and can be attributed to shear stress on the endothelial membrane that contributes to atherosclerosis [Webb and Werring, 2022]. Such chronic microvascular injuries also promote the spread of inflammatory factors through damaged blood-brain barriers as a potential mechanism of HT-associated neurodegeneration [Youwakim and Girouard, 2021]. In the current study, however, CVR amplitude was not significantly altered in the HT group. This can be attributed to the modest sample size and potentially high variability amongst HT individuals. This may also speak to the possibility that CVR amplitude is not the most sensitive CVR marker for HT pathology. There is minimal evidence of sex differences in the CVR impairment.

Diabetes mellitus (DM) and HT are commonly comorbid, and vascular dysfunction has long been known as a part of DM pathology. Thickening of the vascular membrane has been identified as one of the hallmarks of DM, associated with reduced vascular elasticity [Feener and King, 1997; Giordani et al., 2014]. Using transcranial Doppler ultrasound (TCD), CO_2_-based CVR in the middle-cerebral artery was found to be impaired in DM [Kadoi et al., 2003]. However, studies of the effect of DM on CVR in human populations are still scarce. Ivankovic reported TCD-based vascular reactivity reductions [Ivankovic et al., 2013], echoing findings in the rat model. One of the few human MRI studies of DM+HT used the BH challenge to localize the CVR deficit to the occipital lobe [Tchistiakova et al., 2014]. Like HT, DM is also associated with blood-flow impairment [An et al., 2018; Jansen et al., 2016] as well as psychological symptoms, such as depression [Jansen et al., 2016]. Damage of the proximal tubule, involved in the uptake of vascular endothelial growth factor (VEGF) [Petrica et al., 2014], as well as a reduction in the brain-derived neurotrophic factor (BDNF) [Zhen et al., 2013], have been implicated in vascular and cognitive pathologies in DM, respectively. VEGF and BDNF are heavily involved in promoting neuronal survival through neurogenesis and angiogenesis, respectively. In the current study, CVR amplitude was found to be significantly reduced in the frontal, precuneus, posterior-cingulate and pericalcarine regions (**Fig. 6**). To our best knowledge, this is the most extensive and detailed list of brain regions to be reported in association with CVR in DM, and these findings are consistent with regions previously reported to exhibit CVR impairment in HT [Hussein et al., 2020] and DM [Tchistiakova et al., 2014], reflecting the comorbidity in this DM+HT group. The implicated regions are known to exhibit high rates of metabolism [Raichle et al., 2001] and consequently, high blood flow [Chen et al., 2011], rendering them more susceptible to vascular damage. Like in the case of HT, there is minimal evidence of sex differences in the CVR impairment.

### 4.3. Estimation and interpretation of CVR delay

CVR timing has been receiving increasing attention as a marker for detecting vascular abnormalities [Donahue et al., 2015; Leung et al., 2016; Sam et al., 2016; Stickland et al., 2021; Thomas et al., 2014; Thrippleton et al., 2018] Using inspired CO_2_ challenges, Leung et al. uncovered longer CVR delays in sickle-cell disease [Leung et al., 2016]. Furthermore, Holmes et al. demonstrated extended CVR delay as a marker with superior sensitivity to the effects of age and Alzheimer’s disease, even when compared to the long-established CVR amplitude measures [Holmes et al., 2020].

In past literature, the BOLD temporal-lag structure has also been estimated in the resting-state BOLD signal by regressing the low-frequency (∼0.1 Hz) arterial BOLD signal [Tong and Frederick, 2014], venous BOLD signal [Christen et al., 2015; Tong et al., 2018] or global BOLD signal [Amemiya et al., 2020; Mitra et al., 2014]. It is understood that the BOLD signal is an indirect measure of blood traversal, as it reflects variations in both blood volume and blood oxygenation. Moreover, the use of the resting-state global signal as a vascular regressor is not entirely supported in concept or by experimental evidence [Scholvinck et al., 2010], and the arterial signal is not always available due to acquisition coverage limitations, for instance. An alternative is to track the BOLD signal change during a hyperoxic or hypercapnic hyperoxic (i.e. carbogen) gas challenge [Yao et al., 2021]. This latter alternative is easier to administer clinically than blood tagging while eliciting a generally robust BOLD response [Donahue et al., 2015]. Indeed, CVR-delay estimation is more robust when performed for a respiratory challenge than using resting-state data [Gong et al., 2023; Stickland et al., 2021; Zvolanek et al., 2023].

The BH task has previously been used for CVR-delay mapping [Aso et al., 2017; Geranmayeh et al., 2015; Moia et al., 2021]. Moreover, the use of Fourier analysis to elegantly estimate the BOLD-CO_2_ response lag time was previously proposed with the use of a sinusoidal CO_2_ stimulus [Blockley et al., 2011]. It was noted that head motion could negatively affect the accuracy of time-lag estimation using BH BOLD fMRI. Moia et al. demonstrated the advantage of using independent component analysis on multi-echo BOLD data to derive a motion-minimized model-independent BH regressor for CVR estimation [Moia et al., 2021]. The delay times are most commonly estimated as the time shift corresponding to the maximum cross-correlation or most significant linear regression in a GLM between the reference and the BOLD signals, with the reference signal being: (1) the PETCO_2_ recording, (2) the whole-grey-matter (GM) BOLD signal [Niftrik et al., 2016; Tong and Frederick, 2014], and (3) the respiratory variability signal (RVT) [Zvolanek et al., 2023].

In our approach, the CVR delay is estimated from the phase of the filtered BOLD signal spectrum and is equivalent to the time shift mentioned earlier. However, unlike GLM-based approaches, no response model is required in our approach. Moreover, consistent with previous work demonstrating the advantages of voxel-wise sinusoidal BOLD response frequency and phase adjustments [Niftrik et al., 2016], our data-driven Fourier-domain pipeline was able to achieve robust CVR delay estimates. Pinto et al. added higher-frequency harmonics to the original single-frequency sinusoidal regressor and demonstrated improvements in CVR model fits [Pinto et al., 2016]. Equivalently, if needed, our method could easily incorporate the phases of higher-frequency spectral peaks in the delay calculation. Our chosen reference is the respiratory-belt signal, which is largely driven by BH-related variations. This is not equivalent to the RVT reference, which is based on lower-frequency respiratory variations. Our estimated delay embodies not only the transit for the CO_2_ bolus to reach the brain region of interest but also the inherent time-to-peak delay of the CO_2_ response function. The former reflects both systemic blood-flow velocity and transit through the main cerebrovascular arteries, while the latter reflects localized vascular elasticity. It is unfortunately not possible to separate these two quantities unless a deconvolution is performed, presenting its own technical challenges [Atwi et al., 2019; Shams et al., 2022]. The chief alternative reference signal, in the absence of the respiration belt recordings, is the whole-grey-matter BOLD signal [Zvolanek et al., 2023], especially as the global BOLD signal variations during a BH task is largely driven by breathing. The main advantage of the GM-BOLD signal is that no external monitoring equipment is required.

In our work, CVR delay in healthy adults spans the range of 10 - 20 s in the GM. Frontal cortical regions and deep-grey regions exhibit the longest CVR delays, while the temporal and parietal regions tend to exhibit shorter delays. These observations are consistent with the work of Trippleton et al. in which the longest GM CVR delays are in the thalamus and the posterior cortex [Thrippleton et al., 2018]. However, while many publications show maps of CVR delay, few quantify regional differences, limiting our ability for cross-validation. The observed spatial diversity in CVR delay may in part be driven by differences in flow patterns in various cerebrovascular territories, including flow transit time and dispersion among others. The temporal and frontal regions, for instance, are supplied by different arterial offshoots, and borrowing from the arterial-spin labelling literature, arterial transit time from the base of the brain is thought to be longest in the occipital region and shortest in deep GM, with flow dispersion following a similar pattern [Gallichan and Jezzard, 2008]. Thus, we are led to think that vascular anatomy is not the main driver of these regional delay differences, but rather, CVR delay is driven by regional vascular elasticity. This supports the utility of CVR delays as potential early indicators of regional physiological integrity.

Of course, the respiratory-belt reference and PETCO_2_ are not fully synchronized. For instance, we used BH upon exhale, and an increase in the belt signal corresponds to the inhale following the BH. In the period immediately after the exhale, CO_2_ slowly begins to accumulate intravascularly over the 15 s BH period. Exhaled CO_2_ variations in turn lag the BH pattern by approximately 15 s, with the PETCO2 value being measured only at the end of the expiratory plateau, which extends over a 4-6 s period. Thus, our respiratory-belt based CVR delay times are likely much longer than those estimated using PETCO_2_ traces. Nonetheless, the advantages of the respiratory belt are apparent when the quality of PETCO_2_ recordings is insufficient, which can often be the case [Zvolanek et al., 2023]. Moreover, for PETCO_2_ recording, the need for extra equipment and often insertion of a nasal cannula, with a stipulation to breathe through the nose or through a facial mask can render participants uncomfortable.

### 4.4. CVR time lag in diabetes and hypertension

DM is well associated with reduced systemic [Rendell et al., 1989] and cerebral blood flow velocity [Jansen et al., 2016; Novak et al., 2006]. Reports of CVR amplitude deficits have implicated the bilateral occipito-parietal regions [Tchistiakova et al., 2014]. In this work, the main DM-related finding is that CVR delay is a more sensitive marker of diabetes than CVR amplitude, as DM+HT participants exhibited longer CVR delays in more extensive GM regions (**Fig. 9**), and delay is associated with more variables than amplitude (**Fig. 7**). The lengthened CVR delay can in part be attributed to the reduced systemic blood-flow velocity, such as associated with vascular stiffness [Jefferson et al., 2018]. Regional cerebrovascular damage due to such factors as hyperglycemia [Giordani et al., 2014] may also stem from the reduction of the blood-derived neurotropic factor [Zhen et al., 2013], further impairing endothelial repair and survival [Kermani and Hempstead, 2019] in what is potentially a vicious cycle. Such mechanisms of endothelial dysfunction may well lead to regionally dependent CVR-response slowing akin to those of other processes such as Alzheimer’s disease and aging [Peng et al., 2018]. The greater sensitivity of CVR delay than CVR amplitude for detecting the association with disease severity and patient cohort effects may suggest that the timing of a preserved CVR response is more clinically significant than the response amplitude itself.

The superior sensitivity of CVR delay to pathology extended to the HT group. One striking finding in this work is that the CVR delay time is generally shorter in the HT group than in either the CTL or HT+DM group, and this finding applies to a large number of ROIs. In contrast, there was no significant difference in CVR amplitude between the HT and CTL groups (**Fig. 8**), suggesting that a healthy CVR amplitude may belie early endothelial pathology [Webb and Werring, 2022] and that using CVR amplitude alone may lead to missed opportunities for understanding the cerebrovascular mechanisms of HT. While counterintuitive, the finding of reduced CVR delay in HT is consistent with prior ultrasound-based reports of increased blood-flow velocity in the presence of elevated blood pressure [Perret and Sloop, 2000]. Recent work using a spontaneous hypertensive model demonstrated an increase in reactive astrocytes and a reduction in microvascular density grew with HT duration [Li et al., 2021]. These changes may well contribute to a pathological speeding of the CVR response. Notably, hypertension is often comorbid with diabetes mellitus [Ivankovic et al., 2013]. However, it is common for diabetes patients to receive hypertension treatment, and as demonstrated in this work, the effect of DM on CVR delay in the DM+HT group surpasses that of HT alone.

### 4.5. Limitations

Our data-driven robust CVR approximation algorithm successfully estimates the fundamental BHF from BH BOLD data for sinusoidal modelling of CVR for BH paradigms with reasonably similar ‘BH’ and ‘baseline’ periods. However, we recognize that many studies may use highly asymmetrical BH timing paradigms that may not be accurately modelled by a sine-cosine function at the fundamental frequency. This can be easily addressed by adding harmonics into the signal as reported previously for sinusoidal modelling [Pinto et al., 2016].

Likewise, while we use a single most common frequency to characterize the CVR for each participant in this demonstrative study, we can add secondary frequencies in the CVR calculation. Nonetheless, in our own secondary analyses including 3 instead of 1 frequency peak for CVR estimation, the CVR amplitude and timing differences between the groups remained unchanged, demonstrating the robustness of choosing a single “representative” BH CVR frequency.

Finally, our study used a limited number of participants. In particular, our quality-assurance standards resulted in a comparatively small DM+HT sample. In practice, it was challenging to meet our recruitment criteria particularly for the HT and DM+HT groups, resulting in a modest sample size. We hope to replicate and expand on our findings in future studies.

## 5. Conclusion

An adaptive data-driven approach is presented for estimating CVR from BH fMRI data to prevent errors due to subject non-compliance and regional CVR time delay variability. Our frequency-domain-based approach for CVR estimation ensures robustness in estimating the CVR using the BH task and serving as quality control of BH data, without the need for PETCO_2_ recordings. The CVR amplitude is estimated in units of %ΔBOLD directly from the data-driven BHF. Serious deviations from the designed task paradigm were suppressed and thus did not bias the estimated CVR values. The voxelwise CVR time delay is also estimated relative to a meaningful non-brain reference point, such as the ventricles, or an external signal like a respiratory belt recording in this work. Our Fourier-spectrum based approach provides an elegant means to estimate CVR time delay estimations using phase differences relative to the reference signal that depends on the BH task.

Our robust data-driven CVR amplitude and time delay estimation can have a wide range of applications in studying patient populations. We demonstrated our method in the study of diabetes and hypertension. The CVR amplitude was lowest in HT+DM, and HT had a lower CVR amplitude than CTL regionally but the voxelwise comparison did not yield statistical significance. Interestingly, the CVR time delay was far more sensitive than the CVR amplitude to differences across the groups. While HT+DM seems to confer longer CVR delays, HT seems to confer shorter delays than CTL. These are the first MRI-based observations of CVR time delay differences between hypertensive-diabetes patients and healthy controls, demonstrating the feasibility of extracting CVR time delay using BH challenges and the unique clinical value of CVR time delay information.

## Data Availability

Data produced in the present study may be made available only according to terms in the research ethics approval.

## Acknowledgements

We are grateful to the Canadian Institutes of Health Research (CIHR) for funding support.

