## Supplementary material for "Robust estimation of dynamic cerebrovascular reactivity using breath-holding fMRI: application in diabetes and hypertension"

**Table S1. Results of the D'Agostino-Pearson normality test for regional means in each group.** CVR amplitudes are normally distributed in all groups, but time delay values are not when the distributions of mean values for all regions are considered.

| Group | CVR | Time delay |
|---|---|---|
| CTL | Yes | No |
| HT | Yes | No |
| HT+DM | Yes | No |

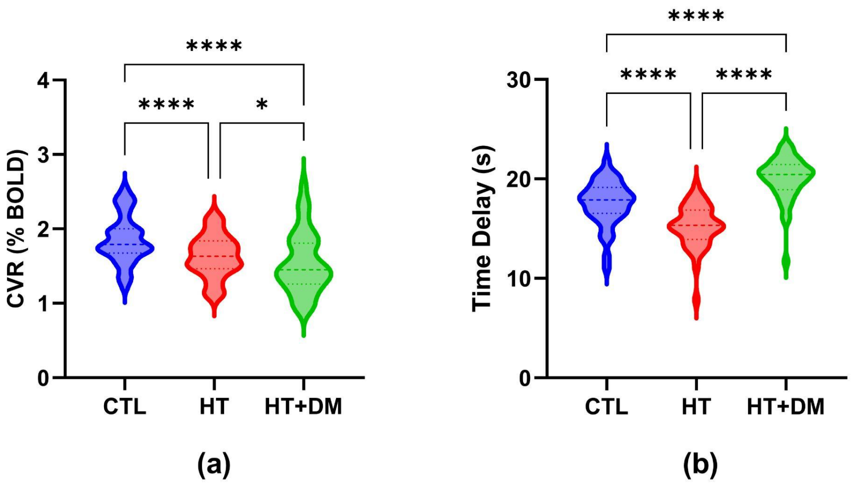


**Figure S1. Mean CVR (a) amplitude and (b) time delay across all cortical ROIs.** The statistically significant differences in CVR amplitude detected by two-way ANOVA and in time delay detected by Friedman's test corrected for multiple comparisons by controlling the false discovery rate using the Benjamini-Hochberg procedure indicated by * ($q < 0.05$) and **** ($q < 0.0001$).
